# Closing the Knowledge Gap: The Importance of Autism Education for Medical Professionals in Kazakhstan, Shaped by Parents’ Experiences

**DOI:** 10.1101/2025.05.25.25328299

**Authors:** Faye Foster, Moldir Tazhibay, Akbota Kanderzhanova, Akbota Tolegenova, Paolo Colet, Valentina Stolyarova, J Gareth Noble

## Abstract

**Background:** In Kazakhstan, families of autistic children face significant barriers in obtaining timely diagnoses and accessing appropriate care. These challenges are shaped by cultural stigma, healthcare system limitations, and insufficient professional training. Understanding parents lived experiences can help inform more effective training for medical professionals.

**Methods:** This qualitative study conducted semi-structured interviews with ten parents of autistic children from both rural and urban areas in Kazakhstan. Participants were recruited through social media outreach, follow-up from a prior survey, and snowball sampling. Interviews were conducted in Kazakh or Russian, transcribed, translated, and analysed thematically.

**Results:** Five key themes emerged: (1) cultural stigma and dismissive attitudes from medical professionals; (2) over-reliance on pharmacological treatments; (3) reliance on informal support networks; (4) limited availability and high cost of autism services; and (5) lack of specialized training among healthcare providers. These barriers contributed to diagnostic delays, emotional distress, and inconsistent quality of care.

**Conclusions:** Parents’ experiences reveal urgent gaps in Kazakhstan’s autism care infrastructure and professional training. Findings highlight the need for culturally adapted training for medical professionals that addresses stigma, improves communication, and promotes evidence-based, non-pharmacological interventions. Integrating parent perspectives into curriculum design can enhance service responsiveness and promote more equitable autism care across the country.

**Lay abstract:** Many parents of autistic children in Kazakhstan face serious challenges when trying to get a diagnosis and access care. These challenges often come from medical professionals who may not be trained to recognize the early signs of autism or who hold negative attitudes. As a result, parents are often blamed for their child’s behaviour, dismissed by doctors, or given limited information about what autism means.

This study interviewed parents from across Kazakhstan to understand their experiences with the healthcare system. Parents shared feelings of frustration, confusion, and emotional distress during the diagnostic process. They also talked about relying on online groups or other parents for support because professional guidance was often unavailable, expensive, or hard to access, especially outside of big cities.

The results showed five major barriers: stigma and blame from professionals, pressure to use medications, limited access to services, poor communication, and a general lack of autism training for doctors. Based on these insights, the study suggests that future training programs in Kazakhstan should include information about autism, communication skills, and support for families. The goal is to help medical professionals better understand autism, reduce stigma, and offer the right kind of help from the beginning.

## Introduction

Autism Spectrum Disorder (ASD) is a complex neurodevelopmental condition that affects communication, behaviour, and social interaction. For the purposes of this article, we will use the term “autism” instead of ASD. While ASD is the medical term, it is rooted in the medical model of disability, whereas “autism” aligns with the positive social model of disability.

Early identification and intervention are widely recognized as key to improving long-term outcomes for autistic individuals and their families. Over the past two decades, there has been growing international emphasis on improving autism awareness, diagnostic accuracy, and access to appropriate services, particularly through the training of medical professionals (World Health Organization, 2023).

### Disparities in Low- and Middle-Income Countries

Despite these advances, significant disparities remain, particularly in low- and middle-income countries (LMICs), where autism awareness and service provision continue to lag. Structural healthcare limitations, combined with cultural stigma, often delay diagnosis and reduce access to evidence-based care. For instance, in many LMICs, healthcare professionals receive little formal education in neurodevelopmental disorders, resulting in misdiagnoses, poor communication with families, and an over-reliance on pharmacological treatments (Zeleke et al., 2021; Duggal et al., 2020). These challenges are evident in Central Asian countries such as Kazakhstan, where health systems face systemic gaps in specialized care capacity (Kosherbayeva et al., 2024; Nukeshtayeva et al., 2022; Foster et al., 2025a, 2025b).

In Kazakhstan and neighbouring countries, the transition from Soviet-era centralized healthcare models to more decentralized systems have left critical shortages in paediatric and neurodevelopmental services, including those required for autism diagnosis and management. One key barrier is the lack of training programs explicitly tailored to autism care, resulting in delayed diagnoses, underutilization of screening tools, and poor integration of autism-specific interventions into routine practice (Kosherbayeva et al., 2024; Somerton et al., 2022).

Compared to upper-middle-income nations such as the United Kingdom and the United States, autism research in Kazakhstan is limited. Somerton et al. (2022) attribute this to a variety of factors, including scepticism about research, difficulties in reaching specialists, and inaccurate or incomplete data reporting by government authorities.

### Autism Diagnosis in Kazakhstan

Existing research is limited, but findings have shown that throughout Kazakhstan, inadequate training leaves medical specialists (psychologists, neurologists, paediatricians) ill-equipped to identify early signs of autism (Somerton et al., 2022; Foster et al., 2025a). These professionals are often unaware of subtle developmental indicators, and the diagnostic process remains misaligned with international standards like DSM-5 and ICD-11. Efforts have been made to adapt validated tools such as the Modified Checklist for Autism in Toddlers, Revised, with Follow-Up (M-CHAT-R/F) to local Kazakh and Russian contexts (Nukeshtayeva et al., 2022). However, the integration of DSM-5 criteria into training frameworks or diagnostic protocols remains limited, leaving ICD-10 as the dominant classification (Kosherbaeva et al., 2024; Kosherbayeva et al., 2023; Kosherbayeva et al., 2024) despite the introduction of ICD-11 by the World Health Organization in 2022.

Research has highlighted challenges in transitioning between diagnostic frameworks. DSM-5 offers a unified ASD diagnosis with severity levels, while ICD-10’s rigid subcategories still dominate in Kazakhstan. These differences, combined with uneven healthcare capacity, exacerbate discrepancies in diagnosis and care when international standards are introduced without proper training and support (Government of the Republic of Kazakhstan, 2021; Nukeshtayeva et al., 2022; Kosherbaeva et al., 2024; Kosherbayeva et al., 2024; Somerton et al., 2022).

Nonetheless, recent efforts aim to improve Kazakhstan’s autism diagnostic system. Initiatives include adapting culturally tailored diagnostic tools (Nukeshtayeva et al., 2022; Gulati et al., 2019), training healthcare professionals in autism awareness (Kosherbaeva et al., 2024; Kosherbayeva et al., 2024), and enhancing intersectoral cooperation among healthcare, education, and social services (Government of the Republic of Kazakhstan, 2021; Kozhageldiyeva et al., 2023). However, these initiatives are in early stages, and the policy framework still lacks a strong focus on autism-specific diagnostics and support (Government of the Republic of Kazakhstan, 2021; Kozhageldiyeva et al., 2023; Kosherbayeva et al., 2025).

Notably, a national “Roadmap for Comprehensive Assistance to Children with Disabilities (2021–2023)” demonstrates progress toward modernizing healthcare and education systems (Government of the Republic of Kazakhstan, 2021; Sakenova et al., 2024; Makoelle, 2020). There has also been an expansion of rehabilitation services and an emphasis on inclusive education frameworks (Government of the Republic of Kazakhstan, 2021; Kozhageldiyeva et al., 2024). However, implementation is uneven. Rural areas continue to face gaps in access to specialized care, infrastructure, and trained professionals (Government of the Republic of Kazakhstan, 2021; Kozhageldiyeva et al., 2023; Zakirova-Engstrand & Yakubova, 2024; Kosherbayeva et al., 2024; Somerton et al., 2022).

To reduce diagnostic delays and improve access, Kazakhstan should expand training programs, integrate autism-specific protocols into rural clinics, and strengthen coordination between healthcare and education sectors. Implementation efforts should prioritize underserved regions. By prioritizing specialized training and improving infrastructure, Kazakhstan can move toward more timely and effective autism care, particularly if medical training curricula are informed by the lived experiences of the families who use these services.

### Specific Gap: Parents’ Perspectives Underrepresented

Parents play a central role in seeking diagnoses and navigating fragmented care systems. Their perspectives reveal gaps in coordination, provider communication, and the accessibility of autism services. However, few studies in Kazakhstan have qualitatively explored parental experiences with autism services, and even fewer have used these perspectives to shape healthcare provider training (Foster et al., 2025b; An et al., 2020; Alibekova et al., 2022). Given parents’ central role, understanding their experiences offers an opportunity to tailor professional education in ways that are culturally sensitive and practically grounded (Boshoff et al., 2019; O’Neill & O’Donnell, 2024).

### Study Aim

In response to this gap, the present study forms part of a broader initiative (funded by Nazarbayev University) that aims to enhance Autism training for medical professionals in Kazakhstan. The overall initiative will focus on closing significant gaps in knowledge and awareness that are necessary for early detection and intervention.

As part of this work, researchers will conduct qualitative interviews with parents of autistic children. These interviews will explore their experiences within the healthcare system, the challenges they may face in obtaining a diagnosis, and the nature of the support they will receive.

The findings from these interviews will be presented in the paper, offering key insights that will help tailor the training program to better meet the needs and experiences of families.

## Methods

The research was conducted using qualitative interviews with parents of children with autism in Kazakhstan. These interviews provided in-depth insights into the lived experiences of families dealing with autism, focusing on their interactions with the healthcare system, the challenges they face in obtaining a diagnosis, and the support they receive.

Recruitment took place through advertising on social media, from parents who had voluntarily left their contact details after completing an earlier survey, and snowball sampling. A total of 10 interviews were conducted with participants from both rural and urban locations in Kazakhstan and were carried out between January and April 2024. By the eighth interview, responses began to repeat, indicating no new information was emerging. The data was rich and detailed, supporting each identified theme. Familiarity with the subject matter allowed for accurate prediction of responses, showing consistent patterns and a coherent narrative. Consequently, data saturation was concluded. To ensure robustness, two additional interviews were conducted, confirming data saturation after the tenth interview. The semi-structured question schedule was designed to collect insights from parents regarding their experiences with medical and health care professionals throughout the diagnosis and treatment of their children with autism. While the study shed light on key challenges, it also identified positive examples of effective strategies and good practices. The questions are categorized as follows:

1. **Initial Interactions**: Understanding parents’ first consultations and the responsiveness of medical professionals to their concerns.
2. **Diagnostic Process**: Exploring communication, support, and clarity of information provided by medical professionals during the diagnosis.
3. **Ongoing Care**: Exploring follow-up care, coordination with other healthcare providers, and accessibility of medical professionals.
4. **Physician-Patient Relationship**: Exploring empathy, understanding, and involvement in decision-making.
5. **Challenges and Improvements**: Identifying challenges faced and suggestions for improving medical professionals support for families of children with autism.

Given the large size of Kazakhstan, the interviews were conducted online to ensure convenience for participants from all regions of the country. Interviews were conducted in Kazakh and/or Russian. The transcripts were initially translated into English, followed by a back-translation performed by a different translator to ensure accuracy and consistency.

The interviews were audio/video recorded to facilitate transcription. Interviews took on average 60 to 90 mins. At the end of each interview, participants were given the opportunity to ask any questions or share their thoughts about their experience of being interviewed.

A process of informed consent was followed after obtaining ethical approval from the Nazarbayev University School of Medicine (NUSOM) Review Ethics Committee (Reference number 2024Feb#02.). This ensured that participants were fully aware of the study’s purpose, procedures, and their rights before agreeing to take part.

### Analysis

The data collected from these interviews were analysed using thematic analysis, a method that involves identifying, analysing, and reporting themes within the data (Braun & Clarke, 2021).

The research team involved in the analysis (FF, MT) conducted thematic analysis in several stages. First, reading the interview transcripts multiple times to fully understand the content. Then generating initial codes by systematically highlighting significant phrases and sentences that captured key aspects of the parents’ experiences, using both inductive and deductive reasoning. These codes were grouped into broader categories based on their similarities and differences.

Next, the categories were reviewed and refined to develop overarching themes and subthemes that captured the main issues faced by the families. For example, the first theme was ‘cultural perceptions,’ with a subtheme of ‘social stigma.’ Each theme was supported by direct quotes from the interviews, providing a rich, detailed account of the parents’ perspectives. This comprehensive thematic analysis offered a nuanced understanding of the parents lived experiences with autism diagnosis.

### Methods: Reflexivity of Researchers

During the research process, a reflexive approach was taken to ensure the quality and rigor of the findings. By continuously reflecting on our own biases, we strived to minimize their impact, making our results accurate and reliable. Our team brings diverse expertise, from educational interventions and mental health to autism education for medical professionals. This allows us to capture a holistic view of the topic, revealing nuances that might otherwise be missed. Our commitment to transparency and rigor enhances the credibility and trustworthiness of our findings. Additionally, by considering our influence, we uphold ethical standards and respect the perspectives and experiences of our participants.

## Results

All participants were married mothers aged between 23 and 35, each with one child diagnosed with autism in Kazakhstan. The children, seven male and three females, ranged in age from 3 to 10 years. Participants were recruited through various channels, including social media, responses to a survey about parents’ experiences with autism diagnosis, and snowball sampling. There was an even mix of rural and urban dwellers among the participants. Some demographic details are withheld to protect anonymity due to the small number of children diagnosed with autism in Kazakhstan.

### Thematic Results

The thematic analysis revealed five key themes related to autism diagnosis and support in Kazakhstan. **Theme 1** highlighted cultural perceptions, with subthemes including reluctance to diagnose, dismissive attitudes, inadequate communication, bias and prejudice, lack of empathy, and negative comments. **Theme 2** focused on the overuse of medications and antipsychotics. **Theme 3** addressed the role of informal support systems. **Theme 4** discussed the limited availability and high cost of services, with a subtheme on the lack of subsidies. Finally, **Theme 5** examined issues within the healthcare system and professional training.

#### Theme 1. Cultural Perceptions and Diagnostic Challenges

Our qualitative research shows that cultural attitudes play a major role in how autism is diagnosed in Kazakhstan. Many people are hesitant to fully accept children with autism, and this resistance is evident in schools and kindergartens, where integration into mainstream classrooms is often discouraged. As one participant (Participant 5) recalled, “The teacher called me and said, ‘Sorry, we can’t take care of your child anymore because he is different. He is not like the other children, so he cannot be here anymore.’”

Parents shared how this lack of acceptance in their communities made them hesitant to seek a formal diagnosis. Participant 2 explained, “Autism is a stigma. People immediately think they are crazy. Parents of children with mental disorders don’t want to tell anyone until the very end that their children have autism.”

This reluctance isn’t just due to social stigma, it was also reflected in the attitudes of medical professionals across different specialties.

#### Subtheme: Reluctance to Diagnose

Parents reported that some medical professionals exhibited a reluctance to diagnose autism. This hesitation was often attributed by the parents to a lack of training or a fear of the stigma associated with the diagnosis. Consequently, parents felt they faced significant delays in obtaining a diagnosis and accessing necessary interventions for their children.

> *“They say, oh, you have such a beautiful child, so smart, so good. Why do you need disability? He will never be able to drive. He will never be able to work in government agencies. You have done so much work. Why are you ruining your child’s fate? Why do you need this?”* Participant 3.

#### Subtheme: Dismissive Attitudes

Many parents reported that medical professionals often didn’t pay attention or dismissed their concerns about their child’s development. Symptoms of autism were frequently attributed to poor parenting or other non-medical factors, rather than being considered as potential indicators of autism. This dismissive attitude contributed to delays in seeking and receiving an accurate diagnosis.

> *“I went to the head of the clinic. When I approached her, she told me most likely it is highly likely that you have a great deal of fault. You are busy with your own affairs and don’t devote time to your child. This is upbringing, she tells me”* Participant 2.

#### Subtheme: Inadequate Communication

Effective communication between medical professionals and parents was often lacking. Professionals didn’t speak Kazakh, tended to use jargon or failed to provide sufficient information about the diagnosis and available support options. This inadequate communication left many parents feeling confused and unsupported during the diagnostic process.

> *“There is not enough information, I had to directly go to internet myself, not just on diagnostics, on treatment methods”* Participant 4

> *“There is little information, especially for Kazakh speakers in general, and there is little information. For example, my child and I first spoke in Kazakh, then, when I started looking for information, watching, I understood that there is nothing at all in Kazakh. That’s why we started in Russian and continued in Russian”.* Participant 6.

#### Subtheme: Bias and Prejudice

Healthcare providers sometimes held biased views about autism, believing that children with autism could not achieve certain developmental milestones or participate in mainstream activities. These prejudices resulted in lower expectations and less encouragement for the child’s development, further marginalizing the affected families.

> *“She [neurologist] described autism, said that he will not love you. Cold-blooded. And for us, my husband, it was just like, you know, well, like someone shot us in the forehead, and we were just so dumbfounded. I cried a lot in her office during the appointment. My husband was angry”* Participant 10.

#### Subtheme: Lack of Empathy and Negative Comments

A notable number of parents experienced a lack of empathy from medical professionals. They reported being treated with indifference or impatience, which made them feel judged and isolated.

> *“In general, there was a feeling that we were not in rehabilitation, but in some kind of prison, you know, serving a sentence, that we were punished, and we are serving a sentence”* Participant 8.

This lack of empathy exacerbated the emotional challenges faced by families dealing with autism. Additionally, some professionals made negative or discouraging comments about the child’s prospects. Such remarks were demoralizing for parents and affected their willingness to seek further help. These negative interactions suggest the need for a more supportive and understanding approach from healthcare providers.

> *“They [rehabilitation staff] told me they wished that [childs name] would become normal, that she would survive. It really pissed me off. I didn’t perceive her as disabled then, really. I perceived her as an ordinary child with some difficulties, and these wishes, I didn’t like them, to put it mildly. I guess that day I realized that my child would never be like others”.* Participant 9

Very few parents reported positive experiences with physicians during the diagnosis and treatment of their children with autism. However, one parent noted that their physician was empathetic, which made a significant difference in their experience.

> *“I like the neuropsychologist. You see, in theory, everything depends on the specialist, no matter who the specialist is. The neuropsychologist we are going to now, I like out of all the ones we have gone to. Basically, many specialists cannot give their all 100%”.* Participant 8.

Additionally, another participant shared a positive experience at an autism centre. Although their interactions at the centre were not with medical professionals, the experience is still worth mentioning as it highlights the importance of supportive environments.

> *“But we took these tests for free in “Assyl Miras”. There is a centre in our city. I went there myself, applied and signed up for diagnostics. It was also very good there, the girls explained and showed everything, all these sites told me where to look for additional information, where these tests are”* participant 4.

### Theme 2. Overuse of Medications/Antipsychotics

Another significant issue raised by parents was treatment approach. Participants noted that doctors frequently prescribed antipsychotics (e.g. risperidone) ostensibly to manage behaviour. Many parents felt these medications were given more to placate them than based on the child’s needs. One parent shared *“All physicians prescribed mountains of medication to calm mothers” (Participant 1).* Another parent echoed this sentiment *“All the doctors, and the other neurologists will at least prescribe some kind of pill. Maybe to calm mothers. That’s it*” (Participant 10).

### While these medications can be effective in reducing irritability and aggression, interviewees highlighted the significant side effects, including sedation, and movement disorders that only exacerbated their concern as parents

> *“They just give antidepressants or these sedatives that bring the child to a state of vegetable”* (Participant 2).

> *“Mostly doctors, they prescribe mountains and mountains of vitamins, pills, yes, examinations, which cost, well, a lot of money, and then you think where to get this money, there, you go, because you think that all this will help”* (Participant 10)

Many felt that these medications were often prescribed in cases where non-pharmacological interventions could have been explored first. However, In Kazakhstan, while there are several non-pharmacological interventions available for individuals with autism, many parents report that these services are either not offered or are prohibitively expensive.

On the occasions when non-pharmacological services were accessed, parents reported that the lack of experience with autism among these professionals negatively impacted the quality of care their children received.

> *“They sent us to the speech therapist. The speech therapist immediately refused us from the clinic, she said I can’t take you because the child needs a neuropsychologist. The child doesn’t sit, he’s hyperactive, I can’t work with him”* (Participant 5).

Participants who took part in the study also felt that doctors in general did not have the skills to help their children.

> *[In reference to ENT physician] “I say, he is non-speaking, he has a diagnosis. She tries to look into his ear, he screams, resists, she freaks out, you know, she freaks out, “What is this? He screams, how am I going to look” and you know, it’s like she’s pouring it all out on me, like we’re deliberately bringing her to this state”* (Participant 7).

### Theme 3. Informal Support

Parents often feel frustrated by the hierarchical healthcare system, which is rooted in the Soviet medical model and frequently limits direct dialogue with medical professionals during the decision-making process. This lack of open communication leaves many parents feeling unheard and unsupported. As a result, they turn to informal networks for help, driven by a sense that the system is not meeting their needs.

One parent shared, “*I started searching and following mommy bloggers on Instagram, with the same diagnosis”* (Participant 3). Another parent echoed this sentiment, saying, “*WhatsApp groups, in all these chats, we mothers get information from each other*” (Participant 7). These networks become a crucial source of support and information for families navigating the complexities of their children’s healthcare.

### Theme 4. Limited Availability and High Cost

All parents articulated their frustration with the unavailability and prolonged waiting times in state clinics.

> *“I initially wanted to make an appointment with a neurologist at my place of residence, for free. But it’s impossible to make an appointment there, there’s such a terrible queue. I couldn’t, I tried to make an appointment for six months. I just wanted to make an appointment with them and I couldn’t make an appointment”* (Participant 4).

> “*I went to a private doctor myself three hundred kilometres from our village, because in our village there are no such specialists”* (Participant 7).

Many parents expressed frustration over the limited availability of non-pharmacological interventions and the prohibitive cost of therapies such as Applied Behaviour Analysis (ABA) and speech therapy. This financial burden makes it challenging for many families to provide consistent and effective care for their children.

> *“I know these treatments exist but contact with them is a different story. There are not enough specialists, the queues are long, and it is too expensive”* (Participant 8).

> *“We work with a private speech therapist, with a defectologist. They also gave us a referral to a rehabilitation center, but since there are few centres, we waited more than a year queue, again, time was lost”.* (Participant 6).

In Kazakhstan, a ‘defectologist’ is a specialist who works with children and adults with developmental disabilities the aim to ‘correct’ or remediate the disability and rehabilitate the individual (45).

Several participants were informed that treatment was not available in their local area and were advised to relocate to the capital city. However, most could not afford the additional costs associated with such a move, including high rents and difficulties finding employment. Despite their desire to access better services, they simply could not manage the financial burden.

> “*The doctor didn’t offer me anything, no options. We got a pediatric neurologist. I went to her, she only recommended that I first inject nootropics with Cortexin. We injected them, and then she said that we need to move to Astana. This is your only way out of this situation, no one here will help you, she said. In Astana, they say, there are all the specialists, but this is also unrealistic for us, for rural residents, how to move to the capital with real estate prices and finding a job is also difficult*” (Participant 8).

#### Subtheme: Lack of Subsidies

Parents reflected that there is a major need for financial assistance and subsidies to make treatments more accessible.

*“We received nothing from the state; we paid for everything” Participant 3*.

> *“I explained the situation, I brought the diagnosis, when they had already made it, I asked to be put on targeted social assistance, but they refused me. Do you know what they told me? That this piece of paper with the diagnosis doesn’t play any role for them. This is wrong”* (Participant 2).

### Theme 5. Healthcare System and Professional Training

Parents spoke of there being a significant shortage of trained specialists, particularly in rural areas. The majority of specialists, including child psychiatrists and developmental paediatricians, work in major cities like as Nur-Sultan and Almaty. Even in these urban regions, parents reported difficulty reaching these specialists and being referred to other medical professionals with different specialties.

> *“They sent me to a neurologist. My neurologist, she said can’t give me any diagnosis, that it’s not their diagnosis, but a mental diagnosis. And she’ll just write me a diagnosis of the central nervous system. She said something with the central nervous system and will send me to neuralgia. That’s all she can do. We need to go to a children’s psychiatric hospital”* (Participant 1).

A prominent theme that emerged from the interviews was the belief that medical professionals lack specialized training in diagnosing and managing autism. This gap in knowledge was a significant concern among participants.

Participants consistently expressed frustration and concern over the perceived lack of specialized training among medical professionals. One parent shared,

> *“It seems to me that we need to raise the level of education in doctors, probably some kind of erudition. In fact, we don’t have many doctors who are now for evidence-based medicine”* (Participant 4).

> *“First, there needs to be early diagnostics. And as soon as there was a suspicion, they should refer to a child psychiatrist, because here anyone can make a diagnosis, speech therapists, psychologists, and neurologists, especially neurologists love it, they will diagnose autism and along the way they start treating, prescribe all sorts of drugs that are ineffective, that are not used anywhere in the world, but drugs, a bunch of everything”* (Participant 9).

## Discussion

This study explored the lived experiences of parents navigating autism diagnosis and care in Kazakhstan. Parents described inconsistent diagnostic experiences, cultural stigma from providers, and limited service options. These issues suggest that autism training for healthcare workers should include both clinical skills and sensitivity to family experiences.

Five major themes emerged:

1. **Cultural stigma and dismissive attitudes** among healthcare providers contribute to diagnostic delays and emotional distress for families.
2. **Over-reliance on pharmacological treatments**, including antipsychotics, often replaces or precedes the use of non-pharmacological, evidence-based therapies.
3. **Informal peer support networks** (e.g., WhatsApp groups, social media) serve as critical, yet unofficial, sources of information and emotional support due to the failure of formal healthcare pathways.
4. **Limited availability and high cost of services**, particularly outside urban centres, exacerbate access disparities and impose a significant financial burden on families.
5. **Deficiencies in healthcare professional training**, including a lack of familiarity with autism diagnostic tools and a lack of empathy, result in inconsistent care and poor patient-provider relationships.

In this discussion, we will explore the study findings and explain how these insights can guide the development of the training courses for medical professionals in Kazakhstan.

### Cultural stigma and dismissive attitudes

The first theme identified in this study was the impact of cultural stigma and negative attitudes among medical professionals, which parents described as a major barrier to obtaining a timely autism diagnosis for their children. A common pattern across interviews was the tendency of healthcare providers to blame mothers for their child’s developmental challenges. Parents reported being dismissed, misunderstood, or accused of poor parenting. These experiences led to delays in assessment, emotional distress, and a loss of trust in professionals.

This aligns with previous research highlighting the widespread experience of stigma among caregivers of children with autism in LMICs. Studies indicate that such stigma can lead to “affiliate stigma,” where parents internalize negative societal views, with harmful consequences for their mental health and engagement with services (Somerton et al., 2022, Malhi et al., 2022).

To address these challenges, training programs must be culturally adapted to Kazakhstan’s unique context. This includes reflective exercises on personal bias, education on neurodiversity models, and recognition of subtle signs of autism. While peer-reviewed evaluations remain limited, descriptive accounts of initiatives such as Project PACE (Anna Freud Centre & AT-Autism, 2025) and HANDS in Autism® (HANDS in Autism®, n.d.) suggest improvements in service engagement. These efforts align with broader research emphasizing the role of culturally grounded, community-based training in reducing stigma and improving care (Hotez et al., 2023).

Parents in this study also expressed dissatisfaction with the way professionals communicated during the diagnostic process. They reported feeling dismissed, not being given enough information, or being spoken to in inaccessible language. These communication issues deepened feelings of frustration and isolation.

Such experiences are supported by earlier studies, which show that poor communication in autism care, such as failure to validate parent concerns or provide accessible explanations, can reduce satisfaction and trust in providers (Legg & Tickle, 2019). Structured training programs like ECHO Autism have been shown to improve professional self-efficacy and family engagement by focusing on neurodiversity-affirming communication and collaborative care planning (Hine et al., 2021, Mazurek et al., 2017).

Parents in this study also shared that they felt unsupported and emotionally dismissed by providers, particularly during diagnosis. These emotional gaps, when combined with stigma, left families feeling isolated. Meta-syntheses of parent experiences in other contexts show that this is not unique to Kazakhstan. Dismissiveness, lack of tailored information, and poor cultural or linguistic fit are well-documented challenges in autism care (Boshoff et al., 2019, Yi et al., 2020, O’Neill & O’Donnell, 2024; Foster et al., 2025b).

While some training programs in LMICs have improved diagnostic skills among professionals, most lack a strong focus on parent-centred empathy or communication. Programs like “Time for Autism” (Dhuga et al., 2022, Gallaher et al., 2023) and the “Embracing Team” (Maia et al., 2016, De Leeuw et al., 2020) demonstrate the value of embedding family perspectives into training design to build emotional intelligence and increase sensitivity among trainees.

Parents reported feeling dismissed and unsupported by healthcare providers. These findings suggest that training should incorporate specific communication techniques, bias reduction strategies, and opportunities for clinicians to hear directly from families. Including parent-led feedback mechanisms may provide valuable, context-specific insights from those directly affected.

### Reliance on Informal Support Networks

A notable finding from this study was that parents frequently turned to informal networks, such as online support groups and peer messaging apps, to seek information and emotional support. These networks often filled the void left by limited professional guidance. Parents shared that these informal platforms were more accessible, responsive, and validating than official healthcare channels. The reliance on peer support reflected both a lack of trust in medical professionals and the limited availability of formal autism services.

This experience is consistent with global patterns, where families navigating autism often report undergoing a “diagnostic odyssey” with minimal guidance from professionals (Lappé et al., 2018). Research suggests that informal support systems can be empowering, but when they become the primary source of information, it indicates systemic deficiencies in healthcare delivery. To address this, healthcare providers must be equipped not only with diagnostic knowledge, but also with communication skills that foster trust and partnership with families. Training curricula that incorporate counselling skills, cultural competence, and relationship-building can help bridge the gap between professional and informal supports.

In addition, Healthcare systems could consider mechanisms to recognize and collaborate with parent-led groups or community advocates, potentially through structured referral systems or advisory partnerships. Doing so would acknowledge their value while helping ensure that families receive accurate, evidence-based guidance.

Parents’ dependence on informal support networks highlights the need for healthcare systems to rebuild trust, enhance communication, and proactively integrate family-centred resources into autism care. Training medical professionals to engage respectfully with both parents and peer advocates can strengthen the bridge between formal and informal systems, especially in under-resourced settings.

### Limited Availability and High Cost of Services

Participants in this study described severe difficulties accessing autism-related services, especially outside major urban centres. Families reported long waiting lists at state-run facilities, and while private services such as Applied Behaviour Analysis (ABA) and speech-language therapy were available, they were often prohibitively expensive. For many parents, the cost of these interventions placed significant financial strain on their families. Rural participants noted that specialized services were rarely available in their communities, and some were advised to move to larger cities to receive appropriate care, which was not a viable option for most.

These findings echo broader research on autism service access in low- and middle-income countries. Ghanouni and Naimpally (2025) observed that uneven infrastructure and urban-centric healthcare planning create substantial barriers for families seeking diagnosis and intervention. Furthermore, studies show that rural families are disproportionately affected by structural inequities, including workforce shortages, transportation barriers, and fragmented referral systems (Pervin et al., 2022).

To address these disparities, policy and training strategies must work together. On the policy side, public investment is needed to decentralize services, subsidize diagnostic and therapeutic costs, and increase the availability of trained providers across all regions. On the training side, general practitioners, paediatricians, and rural health workers must be equipped to deliver basic autism screening, make timely referrals, and provide culturally responsive guidance. Task-sharing models, in which non-specialist providers are trained to deliver community-based autism interventions, offer a scalable, cost-effective approach used in many LMICs (Duggal et al., 2020, Silva et al., 2017).

Parents’ accounts reveal a critical service gap in Kazakhstan’s autism infrastructure. Geographic, financial, and systemic barriers leave many families without access to essential care. By training front-line healthcare providers and investing in decentralization, Kazakhstan can improve early identification and reduce disparities in autism service delivery.

### Over-reliance on pharmacological treatments

Many parents in this study voiced concern over the routine prescription of medications, particularly antipsychotics such as aripiprazole and risperidone, as the first-line response to their children’s behavioural challenges. While some parents acknowledged that medications provided short-term relief for symptoms such as aggression or irritability, most felt uneasy with how readily doctors prescribed them without exploring alternative interventions. Several participants believed that medications were prescribed more for the reassurance of parents than for the child’s actual needs and described a lack of discussion about potential side effects or long-term implications.

This over-reliance on pharmacological treatment has been documented in other studies examining autism care in LMICs. Alsayouf et al. (2021) confirm that while antipsychotics may reduce irritability, they are often used inappropriately or in isolation, without behavioural supports. Parents’ hesitation toward medications reflects a growing body of research emphasizing the importance of evidence-based, non-pharmacological interventions — such as parent training, ABA therapy, and communication-focused strategies — which are often underutilized due to cost, availability, or professional unfamiliarity (Choi et al., 2024, Silva et al., 2017, Zeleke et al., 2021).

In Kazakhstan, limited availability and high cost of non-drug therapies further exacerbate this reliance. Medical professionals may default to medications due to a combination of limited training in behavioural interventions and systemic barriers that restrict access to non-pharmacological care, a pattern observed in several LMICs (Silva et al., 2017; Duggal et al., 2020).

Programs in other LMICs have demonstrated that integrating non-pharmacological interventions into medical training can be both effective and scalable. For example, training healthcare workers to deliver behavioural strategies, parent coaching, and environmental modifications has led to significant improvements in child and family outcomes in countries such as Brazil, Ethiopia, and India (Duggal et al., 2020, Silva et al., 2017).

Parents’ concerns about the widespread use of medications highlight a critical gap in autism care, the lack of access to, and training in, non-pharmacological interventions. Reducing inappropriate medication use in Kazakhstan will require provider education, structural reform, and the inclusion of affordable, evidence-based alternatives within national autism care protocols.

### Deficiencies in healthcare professional training

Across all interviews, parents highlighted a widespread lack of autism-specific training among medical professionals. This deficiency was linked to misdiagnoses, delayed referrals, and dismissive or insensitive treatment during clinical encounters. Participants shared that many physicians did not recognize early signs of autism, were reluctant to discuss the diagnosis, or used outdated terminology. Several reported that professionals deferred to personal opinion or anecdotal knowledge rather than applying established diagnostic frameworks.

This lack of formal training contributed to inconsistent care and a breakdown in trust between families and healthcare providers. Some parents described seeking second or third opinions, often outside of their region, to obtain an accurate diagnosis. Others resorted to self-education or consultations with peers due to a perceived lack of expertise within the healthcare system.

These findings are consistent with existing research from LMICs, which indicates that many physicians, nurses, and allied health professionals receive minimal autism-specific instruction. Training efforts in these contexts tend to focus narrowly on improving diagnostic accuracy or protocol adherence, often overlooking critical competencies like empathy, communication, and family-centred care (Hagag et al., 2024, Kosherbayeva, Kozhageldiyeva, et al., 2024, Silva et al., 2017, Zeleke et al., 2021).

Although some programs have begun integrating family perspectives, these remain the exception. For instance, the “Time for Autism” initiative introduces medical students to families of autistic children, fostering awareness of parental challenges and day-to-day realities (Dhuga et al., 2022, Gallaher et al., 2023). Similarly, the “Embracing Team” model focuses on equipping professionals to support families through the emotional aspects of diagnosis, although it is not yet widely implemented (De Leeuw et al., 2020). To our knowledge, Kazakhstan currently lacks a widely adopted, standardized national framework for autism-specific training in medical education (Somerton et al., 2022). Much of the professional knowledge is self-taught, derived from Russian-language materials, or based on outdated diagnostic classifications such as ICD-10 (Somerton et al., 2022). Without consistent training in current international standards like DSM-5 or ICD-11, healthcare providers may struggle to deliver timely, accurate, and sensitive care. Kazakhstan’s medical professionals face significant training gaps in autism diagnosis, communication, and family engagement. To improve care quality and parent satisfaction, future training initiatives must include not only diagnostic tools, but also emotional support strategies, cultural competence, and parent-informed curriculum design. A standardized, evidence-based framework for autism education is essential to closing this gap.

These five themes demonstrate the ways in which cultural beliefs, economic constraints, and gaps in professional training intersect to create barriers for families navigating autism care. Figure 1 provides a summary of these findings, outlining how each theme contributes to ongoing difficulties in the diagnosis and management of autism in Kazakhstan. The figure also identifies key areas where healthcare training can be strengthened, including strategies to reduce stigma, improve provider-parent communication, and increase the use of appropriate, non-pharmacological interventions.

**Figure 1.**
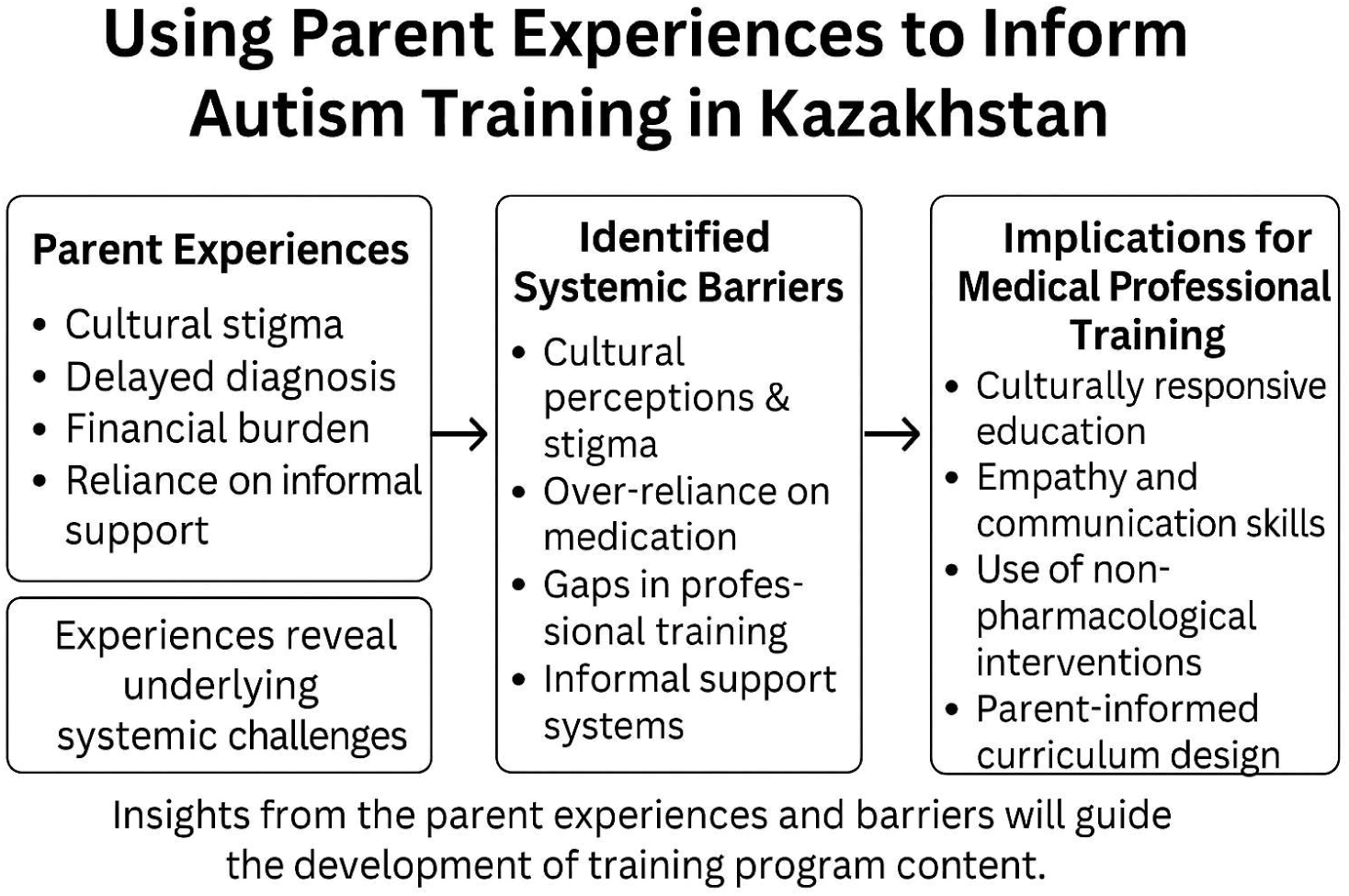
Conceptual summary of key themes from parents’ experiences and their implications for professional training.

### Limitations

The study gives useful insights into Kazakhstani parents’ experiences with autistic children, showing substantial hurdles in their children’s route to diagnosis. However, it has limitations, including a small sample size of 10 interviews, possible selection bias, and a lack of statistical rigor caused by subjective reports. The study’s cultural background may also influence its application to other nations with varying healthcare methods and public views.

Furthermore, the research focuses entirely on parents’ experiences, leaving out the viewpoints of medical professionals and persons with autism. The research also documents a particular period in time, which may be influenced by changes in healthcare legislation or society views. Addressing these limitations in future study might contribute to a more comprehensive knowledge of the issues that families and medical professionals encounter while treating autism in Kazakhstan.

### Conclusion

This study explored the everyday experiences of parents raising autistic children in Kazakhstan, with the aim of informing the development of medical training programs. Interviews highlighted persistent barriers to timely diagnosis and effective care, including cultural stigma, over-reliance on medication, limited professional empathy, restricted access to services, and gaps in provider training.

Although recent policy developments and academic literature point to progress in autism care, parents’ accounts reveal more entrenched and systemic challenges. Their experiences suggest a disconnect between official reforms and the day-to-day realities faced by families, particularly in relation to professional communication, emotional support during diagnosis, and access to appropriate services, especially outside urban centres.

Including parents’ perspectives in medical education offers a practical way to address these issues. Doing so can support the development of a healthcare system that is more responsive to families’ needs, with improved diagnostic accuracy, greater trust between parents and professionals, and more equitable access to evidence-based care, regardless of location or background.

We recommend that training programs for medical professionals in Kazakhstan:

1. Address stigma and bias explicitly,
2. Promote non-pharmacological, family-centred care,
3. Enhance communication and empathy skills,
4. Include practical solutions for service access in low-resource areas
5. Integrate parental voices directly into curricula design.

Making these changes, along with improvements in the healthcare system, could help doctors diagnose autism earlier, ease stress for parents, and give children better chances for support across Kazakhstan.

## Data Availability

The data that support the findings of this study are available from the corresponding author upon reasonable request.

## Declaration of conflicting interests

The author(s) declared no potential conflicts of interest with respect to the research, authorship, and/or publication of this article.

## Funding

This work was supported by Nazarbayev University (Faculty-Development Competitive Research Grants Program, 2023–2025, Project #20122022FD4132).

## Author Contributions

**Faye Foster:** Conceptualization, Methodology, Investigation, Writing – Original Draft, Supervision.

**Moldir Tazhibay:** Investigation, Writing – Review & Editing.

**Akbota Kanderzhanova:** Investigation, Writing – Review & Editing.

**Akbota Tolegenova:** Investigation, Writing – Review & Editing.

**Paolo Colet:** Writing – Review & Editing, Supervision.

**Valentina Stolyarova:** Writing – Review & Editing.

**J. Gareth Noble:** Conceptualization, Writing – Review & Editing, Supervision.

## Notes

### Competing Interest Statement

The authors have declared no competing interest.

### Funding Statement

This study was funded by Nazarbayev University Faculty Development Competitive Research Grants Programme 2023 to 2025

### Author Declarations

Ethical approval was granted Nazarbayev University School of Medicine Research Ethics Committee. Reference number 2024Feb#02, Nazarbayev University.

